# Geographic variation in work disability burden: Identifying hotspots, coldspots, and associated sociodemographic factors

**DOI:** 10.1101/2020.12.08.20246181

**Authors:** Tyler J Lane, Alex Collie

## Abstract

**Objectives:** To identify geographic hotspots and coldspots of work disability burden and associated sociodemographic factors in Australia.

**Methods:** Using Australian workers’ compensation and census data, we calculated weeks of compensated time off work per 1,000 labour force at Statistical Area Level 4, an indicator of work disability burden. Records included all claims with at least one day of compensated time off work lodged between 2010 and 2015. Work disability burden was z-transformed by state and mapped across Australia. Statistical Areas ≥ 1.5 standard deviations from the state/territory mean were considered hotspots and coldspots. We tested several sociodemographic factors as predictors of work disability burden.

**Results:** Work disability burden hotspots were concentrated in lower socioeconomic suburbs and exurbs of state capitals, plus several regional areas. Coldspots were primarily in wealthy central urban and suburban areas. Factors associated with greater work disability burden include socioeconomic disadvantage, rurality, lower labour force participation, higher unemployment, and more people with core activity limitations, aged 65+ years, and aged 65+ but fewer foreign-born.

**Conclusions:** Work disability burden is unequally distributed across Australia and strongly influenced by sociodemographic factors. The findings can guide more efficient allocation of resources for primary and secondary work disability prevention and rehabilitation.

## INTRODUCTION

Work disability occurs when an injury, illness or medical condition restricts the ability of a person to engage in paid employment. Extended periods of work disability are common and can have substantial financial, health and social impacts on the worker.^1,2^ The presence and duration of work disability is now understood to be a product of multiple features of the person and their environment, incorporating social, cultural, health-related, and psychological factors.^2^ These contextual factors vary geospatially. Accordingly, work disability burden varies by location, though there are relatively few studies on this topic.^3^

Injured workers tend to spend more time off work in socioeconomically disadvantaged areas where more people are off work and there are larger populations of ethnic minorities.^4,5^ Rates of work-related injury vary across industry and occupation,^6,7^ which are often geographically concentrated. And while the evidence on rurality is mixed, or suggests at the very least a highly complex dynamic,^8^ people living in rural areas generally have longer work disability duration.^3^

Areas in which multiple of these factors are apparent may be geographic hotspots of work disability, which may warrant a more efficient and targeted reallocation of preventive and rehabilitation resources.^9^ One example of a community approach are the Centers of Occupational Health and Education in Washington state, a community-based occupational healthcare intervention that aims to minimise work disability with secondary health services to improve care coordination and training, incentivise best practice, and engage a broad range of stakeholders. Evaluations suggest these centres reduced both disability duration and the likelihood of long-term disability.^10^

In this paper, we sought to identify hotspots and coldspots of work disability burden in Australia and determine whether it is associated with several sociodemographic factors. There is substantial potential to reduce Australia’s work disability burden and its associated costs. In 2012/13, work injury cost and estimated $61.8 billion AUD, equivalent to 4.1% of Gross Domestic Product.^11^ Even small reductions in work disability could result in substantial savings in addition to the numerous health, social, familial, psychological, productivity, and community benefits.^12^

## METHODS

### Setting

In Australia, workers’ compensation is managed by states and territories. Federal government employees are covered by a Commonwealth scheme, Comcare, which also regulates self-insured interstate employers.^13^ Each system is cause-based, meaning compensation is contingent on establishing that injury or illness is attributable to work.^14^ In 2014, these schemes covered 10.8 million workers, or 94% of the labour force.^15^

### Work disability burden

We calculated the number of compensated weeks off work per 1,000 labour force as an indicator of work disability. Compensation data were extracted from the *National Data Set for Compensation-based Statistics*, a minimum administrative dataset that combines data from each of Australia’s nine major workers’ compensation systems.^16^ We previously used a similar measure to compare work disability burden among Australian first responders,^17^ though there are several caveats. Cumulative compensated time off work underestimates actual time off work^18^ and benefit cessation does not necessarily mark the end of disability duration.^1,18^ Nevertheless, it is considered the most accurate measure that can be derived from administrative claims data.^18^ To account for extreme outliers, individual claims were capped at five years of compensated time off work. The labour force denominator includes all workers, regardless of compensation coverage.^19^ The proportion of uncovered workers likely varies regionally and may be a source of systematic error. While populations change over time, labour force estimates at Statistical Area Level 4 are not available for every year. We used 2015, the only year for which we could find data that falls within our study period.

### Geospatial units

Using residential postcodes, we aggregated claims at Statistical Area Level 4, which is the smallest level for which the Australian Bureau of Statistics produces labour force data and relevant sociodemographic data, and the largest of the Australian Statistical Geography Standards regions below state/territory. Statistical Areas at Level 4 are constructed around labour markets and usually have populations over 100,000.^20^

### Sociodemographic indicators

The Australian census captures a number of sociodemographic factors that have previously been associated work disability burden^4,5^ that are also available at Statistical Area Level 4. These include labour force participation and unemployment rates,^19^ median personal income,^21^ and proportions of foreign-born residents,^22^ needing assistance with core activities, and aged 65+ years (retiree proxy).^23^ We also included proportion with non-proficient English^22^ as a proxy for newly-arrived foreign residents. Other data were aggregated from smaller units. We converted remoteness rankings^24^ to a rurality indicator by scoring postcodes 1 to 5 (from “Major Cities of Australia” to “Very Remote Australia”) and calculating the Statistical Area mean. Socioeconomic advantage and disadvantage is measured with separate variables, calculated as the proportion of postcodes within a Statistical Area that fall within the top and bottom quintiles of the Indicators of Relative Socioeconomic Advantage and Disadvantage.^25^ Each factor was taken from the 2011 Census.

### Inclusion and exclusion criteria

Records were limited to accepted workers’ compensation claims with at least one day of compensated time off work that were lodged between 2010 and 2015. Records are updated for six years after initial recording. At the time of writing, these were updated up to July 2017, providing a minimum of 1.5 years of follow-up. Records were excluded if they did not have postcode information, or the postcode could not be linked to a Statistical Area. The Australian Capital Territory was excluded because it consists of a single Statistical Area, which would not allow scaling that was necessary to account for state fixed-effects (see next section).

### Analysis

Descriptive analyses summarise claims by state and territory with counts, rate per 1,000 labour force, mean disability duration in weeks, work disability burden, and proportion compensated by a different system from the state/territory of residence. This also summarises the Australian Capital Territory or claims that were missing postcode data. We converted work disability burden and sociodemographic data to z-scores. Each was scaled by state and territory, to account for known differences in disability durations between Australia’s compensation systems, likely due to system factors,^26,27^ and known sociodemographic differences between states.^28,29^ This zeroes the means and scales differences between Statistical Areas by standard deviation. Of note, this is not the overall state/territory mean, but the mean of Statistical Areas within states and territories. Scaling also standardises associations for sociodemographic factors, making them directly comparable. We define hotspots and coldspots as Statistical Areas that are at least one standard deviation away from the state/territory mean. Assuming a normal distribution, -/+ one standard deviation is roughly equivalent to the 32^nd^ and 68^th^ percentiles, while -/+ 2 standard deviations is roughly equivalent to the 5^th^ and 95^th^ percentiles. Using geospatial data from the Australian Bureau of Statistics,^20^ we mapped these scaled distributions.

To test associations between sociodemographic factors and work disability burden, we analysed each in separate univariate linear regressions. Multivariable regressions were impractical due to the small number of cases (86 Statistical Areas) and a high degree of multicollinearity (most Variance Inflation Factors ≥ 2.5; see Table 2).

#### Statistical software and data availability

We conducted analyses in R^30^ using RStudio^31^ with the following statistical packages: *broom*.*mixed*,^32^ *janitor*,^33^ *lubridate*,^34^ *magrittr*,^35^ *readxl*,^36^ *see*,^37^ *sf*,^38^ *tidyverse*,^39^ *tmap*,^40^ and *zoo*.^41^ Aggregate data and analytical code are archived on a public repository.^42^ Case-level data are not available because they contain personal identifiers and are sensitive, though we also provide cleaning code to demonstrate how we aggregated data.

## RESULTS

### Descriptives

State and territory work disability profiles are summarised in Table 1. N = 1,010,022 accepted time loss claims were lodged in the study period, of which 0.8% (N = 7,731) are from the Australian Capital Territory and 8.2% (N = 82,367) were missing postcode data and could not be linked to a Statistical Area, leaving N = 919,924 analysable records. Though there are large differences in claim rates and mean compensated weeks between states and territories, work disability burden was relatively similar. The vast majority of claims (96.1%) were compensated by the state or territory in which the injured worker resided. Mean disability duration was longest among claims that could not be linked to a Statistical Area, suggesting that postcode data are not missing at random and the work disability burden measure is an underestimate.

**Table 1.** Summary of state and territory work disability burden for all time loss claims (≥1 day) lodged between 2010 and 2015.

| State/territory | Time loss claims<br>(column %) | % of included<br>claims | Claim rate per<br>1,000 labour<br>force | Mean disability<br>duration<br>(weeks) | Compensated<br>weeks/1,000<br>labour force<br>(work disability<br>burden) | Under state/territory<br>scheme* (row %) |
| --- | --- | --- | --- | --- | --- | --- |
| New South Wales | 350,590 (34.7%) | 38.1% | 96.0 | 13.5 | 1,294 | 337,291 (96.2%) |
| Victoria | 149,126 (14.8%) | 16.2% | 50.3 | 24.1 | 1,211 | 140,134 (94.0%) |
| Queensland | 256,835 (25.4%) | 27.9% | 110.2 | 9.3 | 1,021 | 249,388 (97.1%) |
| South Australia | 49,800 (4.9%) | 5.4% | 61.8 | 20.9 | 1,291 | 47,661 (95.7%) |
| Western Australia | 80,217 (7.9%) | 8.7% | 60.4 | 15.4 | 928 | 77,733 (96.9%) |
| Tasmania | 23,800 (2.4%) | 2.6% | 99.2 | 13.3 | 1,321 | 22,863 (96.1%) |
| Northern Territory | 9,556 (0.9%) | 1.0% | 72.0 | 14.7 | 1,059 | 9,278 (97.1%) |
| **Total** | **919,924 (91.1%)** | **100.0%** | **80.3** | **14.6** | **1,172** | **884,348 (96.1%)** |
| *Excluded* |  |  |  |  |  |  |
| *Australian Capital Territory* | *7,731 (0.8%)* | - | - | *21.6* | - | *2,342 (30.3%)* |
| *Missing postcode* | *82,367 (8.2%)* | - | - | *24.6* | - | - |

**\*Not compensated by a system different from the state/territory in which they live or Comcare**

### Work disability burden hotspots

Hotspots and coldspots of work disability burden are mapped in Figure 1. Hotspots and coldspots more than a standard deviation away from state or territory mean are listed in Table 2; a full table of raw and scaled work disability burdens by Statistical Area can be found in Supplementary Table 1. A Shapiro-Wilks test of normality suggests the distribution is non-normal, though it is barely significant (W = 0.97, p = 0.048). Maximum work disability burden z-scores are two standard deviations in either direction, suggesting no substantial outliers. The histogram in Figure 1 suggests a relatively normal distribution, though fat-tailed.

**Figure 1.**
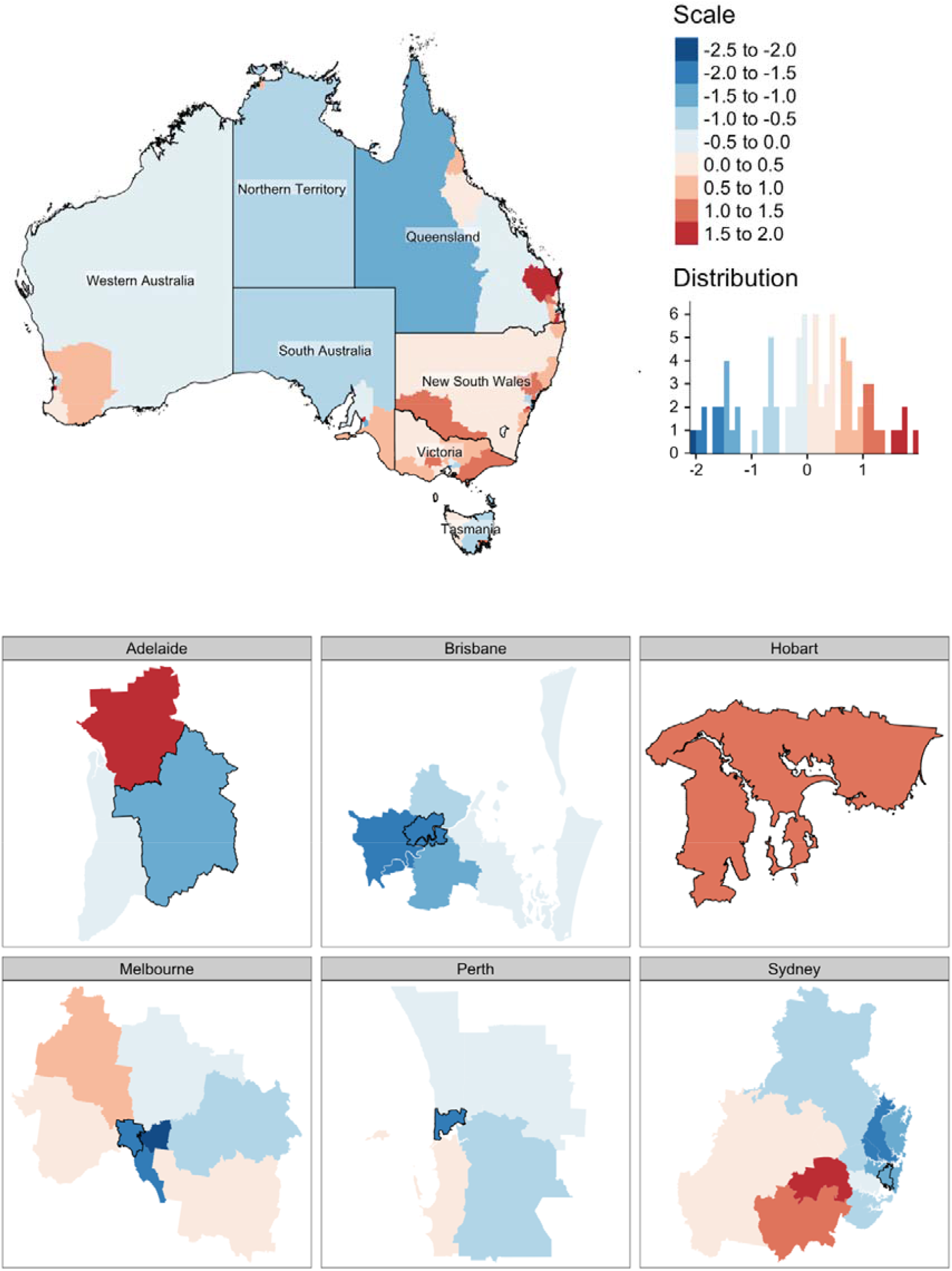
Mapping of scaled work disability between 2010-2015, where blue denotes below state average and red above state average, nationally and by state capitals; Statistical Areas with a black border in city maps are designed central/inner areas

**Table 2.** Top ten hotspots and bottom ten coldspots of work disability burden*.

| State | Statistical Area (#) | Work disability burden† |  |
| --- | --- | --- | --- |
|  |  | Compensated weeks per 1,000 labour force | Z-score scaled by state/territory |
| Hotspots of greater work disability burden (≥ 1 standard deviation) |  |  |  |
| Queensland | Logan - Beaudesert (311) | 1668 | 1.94 |
| South Australia | Adelaide - North (402) | 1827 | 1.71 |
| Western Australia | Mandurah (502) | 1852 | 1.71 |
| Queensland | Wide Bay (319) | 1583 | 1.68 |
| New South Wales | Sydney - South West (127) | 2228 | 1.56 |
| New South Wales | Hunter Valley excluding Newcastle (106) | 2101 | 1.33 |
| Tasmania | Hobart (601) | 1410 | 1.25 |
| New South Wales | Murray (109) | 2024 | 1.19 |
| New South Wales | Central Coast (102) | 2011 | 1.16 |
| Queensland | Moreton Bay - North (313) | 1401 | 1.15 |
| New South Wales | Sydney - Outer South West (123) | 1966 | 1.08 |
| Victoria | Ballarat (201) | 1798 | 1.07 |
| Victoria | Latrobe - Gippsland (205) | 1785 | 1.04 |
| Coldspots of less work disability burden (≤ -1 standard deviation) |  |  |  |
| Queensland | Brisbane - South (303) | 595 | -1.21 |
| Queensland | Queensland - Outback (315) | 571 | -1.28 |
| New South Wales | Sydney - Inner West (120) | 651 | -1.37 |
| New South Wales | Sydney - Northern Beaches (122) | 614 | -1.43 |
| South Australia | Adelaide - Central and Hills (401) | 758 | -1.47 |
| New South Wales | Sydney - City and Inner South (117) | 588 | -1.48 |
| New South Wales | Sydney - Eastern Suburbs (118) | 582 | -1.49 |
| Queensland | Brisbane Inner City (305) | 494 | -1.51 |
| Queensland | Brisbane - West (304) | 476 | -1.56 |
| Victoria | Melbourne - Inner South (208) | 615 | -1.68 |
| New South Wales | Sydney - Ryde (126) | 474 | -1.69 |
| Western Australia | Perth - Inner (503) | 161 | -1.84 |
| New South Wales | Sydney - North Sydney and Hornsby (121) | 384 | -1.86 |
| Victoria | Melbourne - Inner (206) | 480 | -2.00 |
| Victoria | Melbourne - Inner East (207) | 459 | -2.04 |

\* Full table available with archived data and code on Bridges
† Compensated weeks of working time loss per 1,000 labour force, claims lodged 2010-2015

Hotspots (red) are concentrated in more deprived and suburban/exurban areas of state capital cities, including Adelaide - North in South Australia; Mandurah, an exurb of Perth in Western Australia; South West Sydney (better known as the Western Suburbs) in New South Wales; and Logan – Beaudesert in Queensland. The Hunter Valley region around New South Wales’ second-largest city, Newcastle, is also a hotspot. Other hotspots are regional, including Wide Bay and Moreton Bay - North in Queensland, and Murray and Central Coast in New South Wales.

Coldspots (blue) are concentrated in wealthier central areas and suburbs of state capitals. Not one of the top ten coldspots is outside of Melbourne, Sydney (including the wealthy Eastern Suburbs and north shore areas of Ryde and North Sydney and Hornsby), Perth, or Brisbane. To a lesser extent, the same pattern is visible in Adelaide – Central and Hills (z-score: −1.47).

### Sociodemographic indicators and work disability burden

Geospatial distributions of sociodemographic indicators are mapped in Supplementary Figures 1 through 10. All sociodemographic factors except proportion with non-proficient English were significantly associated with work disability burden, as illustrated in Figure 2 and summarised in Table 3. Most associations were in the expected direction: work disability was lower where there was greater socioeconomic advantage and higher where more people were out of work, more people had core activity limitations, and the area was more rural. The strongest associations were observed with the concentration of disadvantaged and advantaged quintiles. For every standard deviation increase in the proportion of postcodes from the most disadvantaged quintiles, work disability burden increased by 0.67 standard deviations (95% CI: 0.51 to 0.83), explaining 45% of the variance, while the most advantaged quintile decreased the burden by −0.73 (95% CI: −0.88 to −0.58), explaining 53% of the variance. The proportion of residents with core activity limitations had a similarly large association, where for every increase in the standard deviation, work disability burden increased by 0.67 standard deviations (95% CI: 0.50 to 0.83), explaining 44% of the variance.

**Table 3.**
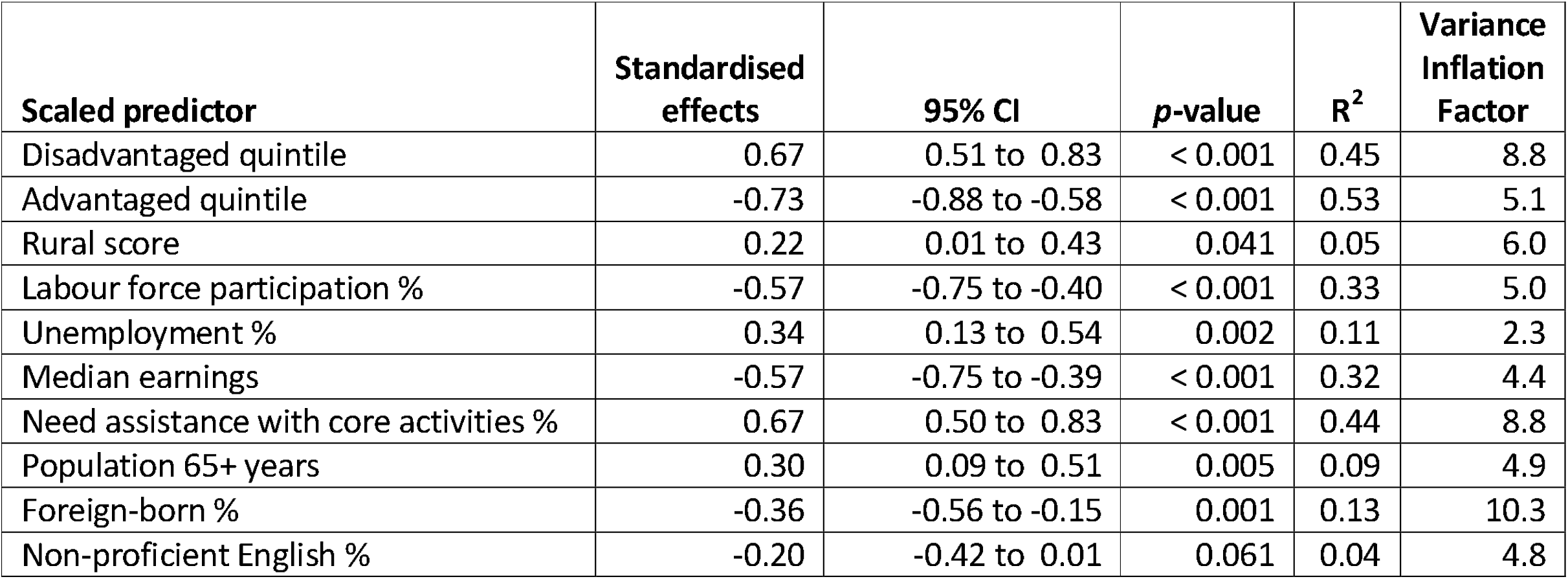
Univariate associations between socioeconomic/demographic factors and work disability burden, scaled for comparability, plus Variance Inflation Factor

The proportion of foreign-born residents was associated with reduced work disability burden (−0.36, −0.56 to −0.15). As this result was unexpected, we examined whether proportion of foreign-born was associated with other sociodemographic variables and whether statistically adjusting for them individually affected the statistical association. All sociodemographic factors were significantly associated with the proportion of foreign-born residents at p < .001, with the exception of unemployment rate (p = 0.299). Areas with more foreign-born residents were more socioeconomically advantaged, less rural, had higher labour force participation and median earnings, with fewer limitations with core activities and older people. When each of the significant factors was added to the foreign-born regression model, the association between proportion of foreign-born residents and work disability burden attenuated to non-significance (p > .05) in all but two cases (population 65+, rural score). These analyses are summarised in Supplementary figures 11 and 12.

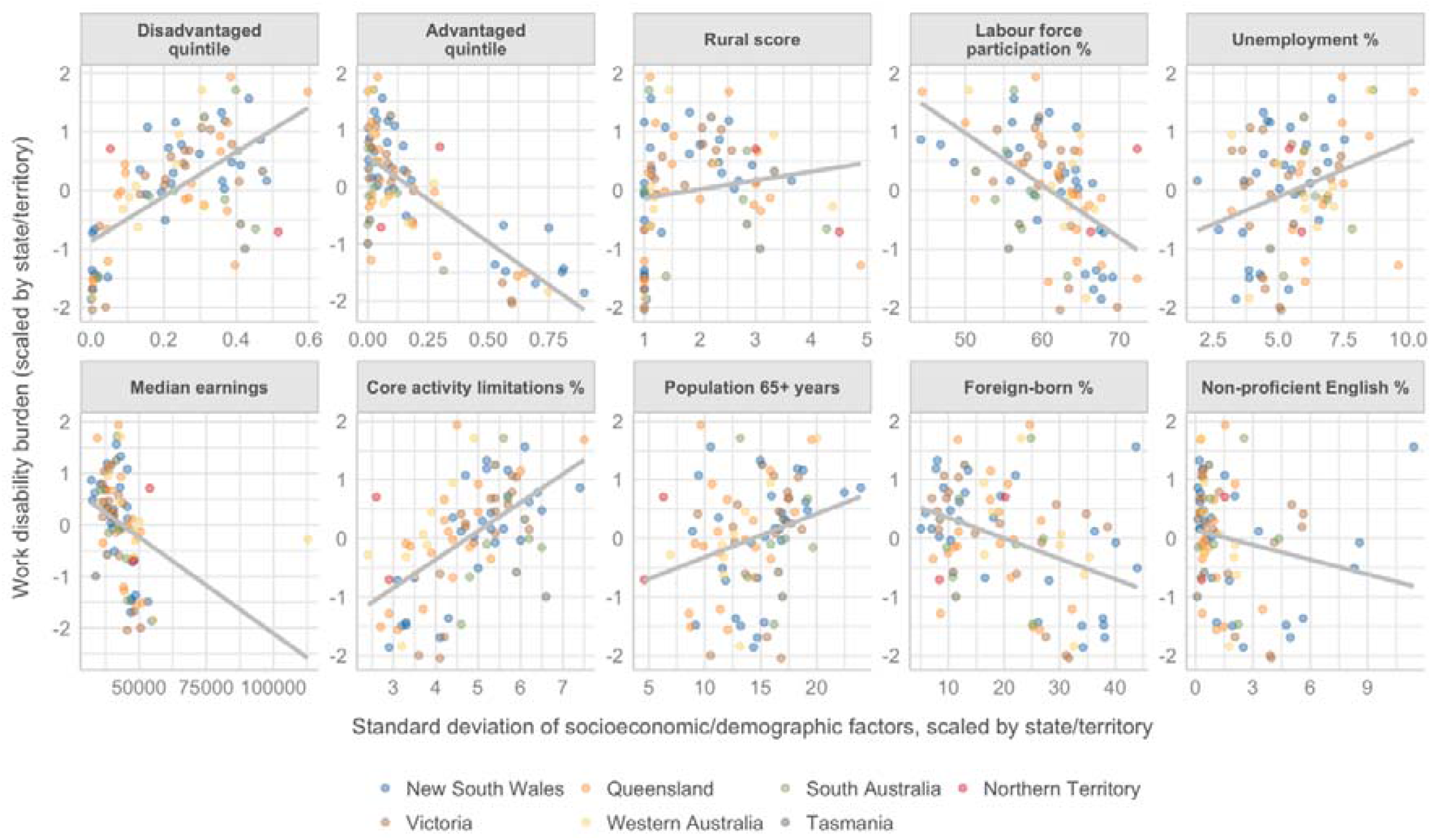

## DISCUSSION

The findings suggest that hotspots of greater work disability burden are concentrated in socioeconomically deprived suburban and exurban areas of the major cities, plus a few regional areas, while coldspots are predominantly socioeconomically-advantaged city centres and their wealthy suburbs. Burden was extremely unequally distributed. For instance, the Statistical Area with greatest burden in Western Australia was Mandurah with 1,852 compensated weeks/1,000 labour force, 11.5 times greater than the Statistical Area with the least burden, inner Perth at 161 weeks/1,000 labour force. This was the most extreme difference, though within other states the differences in work disability burden between hotspots and coldspots was still high at about 3-5:1 (see Table 2).

In line with previous research,^4,5,43^ we found that work disability burden is to a large extent socially-determined. Occupation is a strongly associated with socioeconomic status,^44^ and each carries its own unique set of inherent exposures^6^ and job demand risks.^7^ The nature of work also influences how debilitating an injury is; a physical condition is much more likely to inhibit manual labour than office work.^43^ Education can also determine how well workers can navigate workplace risks, whether through comprehension of safety protocols or opportunities to advance to less risky roles.^43,45^ Socioeconomic status is a strong predictor of general health,^46^ which can moderate the effect of work injury.^47^ Similarly, one of the strongest associations we found was the proportion of residents needing assistance with core activities.

Rurality was associated with higher work disability burden, but only explained 8% of the variance. This was not unexpected as the current evidence on urban/rural differences is mixed, if leaning toward greater burden in rural areas.^3^ Yet there remain a number of indicators to suggest rural Australians should have substantially greater work disability. Rural areas have higher rates of disease, likely exacerbated by lower levels education, greater socioeconomic disadvantage, and poorer access to healthcare services.^48^ Rural areas also have more people employed in higher-risk industries.^8,49^ Indigenous Australians, by any measure the country’s most dispossessed people, also have greater representation in rural areas.^48^ While a different cultural context, areas with more disposed ethnic minorities in the US has previously shown longer disability durations where there are more dispossessed ethnic minorities.^4^ Sociocultural drivers may account for some of the mixed evidence of rurality. Rural residents may be more resilient, which may make them more likely to stay at work and off compensation, while urban residents have easier access to legal services, which is known to delay return to work.^8^ However, rural industries may be underrepresented in workers’ compensation, which could lead to underestimates in these data. For instance, 99% of the public administration and safety workforce is entitled to compensation, compared to only 64% of agriculture, forestry, and fisheries (50).

There is a necessary distinction to be made between work injury and workers’ compensation claims. In this study, we only had records of the latter. Not every injury is compensated,^15^ and even those injuries that become a claim are not compensated for the full duration of time off work.^18^ Individual earnings are one factor. Higher earners are incentivised to exit compensation more quickly^50,51^ due to wage replacement caps, usually set around twice state average earnings,^52^ that reduce the proportion of pre-injury earnings that are compensated. This may account for some of the inverse relationship between higher socioeconomic indicators and reduced work disability burden. Higher unemployment rates may make it harder to leave workers’ compensation since there are fewer alternative employment opportunities.^4,5^ It may also be more socially acceptable to lodge a claim and take time off work where more people are already out of work.^4^

Work disability burdens were lower where the foreign-born share of the population was larger. However, these areas were also more socioeconomically advantaged and less rural, and had greater labour force participation, fewer older people, and people with core limitations. Further, the association between foreign-born populations and work disability attenuated with adjustment for most of these factors. Australia’s emphasis on skills-based immigration means foreign-born residents skew towards the higher-educated, entailing both less risky work and more ability to work safely.^53^ Correspondingly, foreign-born Australians are injured at work (33.0/1,000 population) at one-third the rate of native-born Australians (47.8/1,000 population).^54^ The proportion with non-proficient English was not significantly associated with disability duration, though this should not be interpreted as evidence of no association. With a mean of 1.7% (SD: 1.7), non-proficient English speakers are probably too small a group to meaningfully influence community work disability burdens.

Australia’s social systems tasked with addressing work injury and work disability, including occupational health and safety and workers’ compensation, are resource-constrained. The broad implications of our findings are that addressing vast inequalities in work disability burden, and to reduce work disability at a population level, will require a redistribution of resources. However, modifications to these systems are insufficient to fully address inequalities. Broader coordinated action targeting the social determinants of work disability such as improved education and healthcare are also necessary, though this is a long-term proposition.

### Strengths and limitations

This study has a number of strengths. We used claims data from workers’ compensation systems with near-universal coverage of the Australian workforce. These were combined with sociodemographic census data. Analyses were scaled by state/territory to adjust for system-level differences and allow direct comparisons of the magnitude of associations.

There are also several limitations. Work disability burden calculations may be systematically biased due the variations in the proportion of the labour force who are covered by compensation systems. Around 8% of records were missing postcode data and could not be mapped to a Statistical Areas. These also had the longest disability duration, meaning the work disability burden was a substantial underestimate. The small number of Statistical Areas and high level of multicollinearity rendered multivariable analysis impractical.

## CONCLUSIONS

Geographic variability in work disability is strongly socially-determined. The differences in burden between hotspots and coldspots is stark, though this highlights great potential to reduce burden through the reallocation of resources. Hotspots of work disability burden correspond to suburban, exurban, and regional areas with greater socioeconomic disadvantage, while coldspots are concentrated in central areas of major capital cities and wealthy suburban areas. In addition to socioeconomic factors, rurality and having more people out of work and who need assistance with core activities are associated with greater work disability, though it is lower where there are more foreign-born workers. The findings can be used to more efficiently allocate resources to areas with disproportionately high work disability burden, though it will be necessary to address the social determinants of work disability to achieve population-level reductions.

## Data Availability

Aggregated data and code are available on a Bridges repository. While we do not provide identifiable raw case-level data, we have provided our cleaning Rmarkdown file to demonstrate the process. Where we do not own the data such as state and territory shape files, we have provided a link to the original source in the Rmarkdown file chunk.

https://doi.org/10.26180/5f7f8f8f5df60

## Supplementary figures

**Supplementary figure 1.**
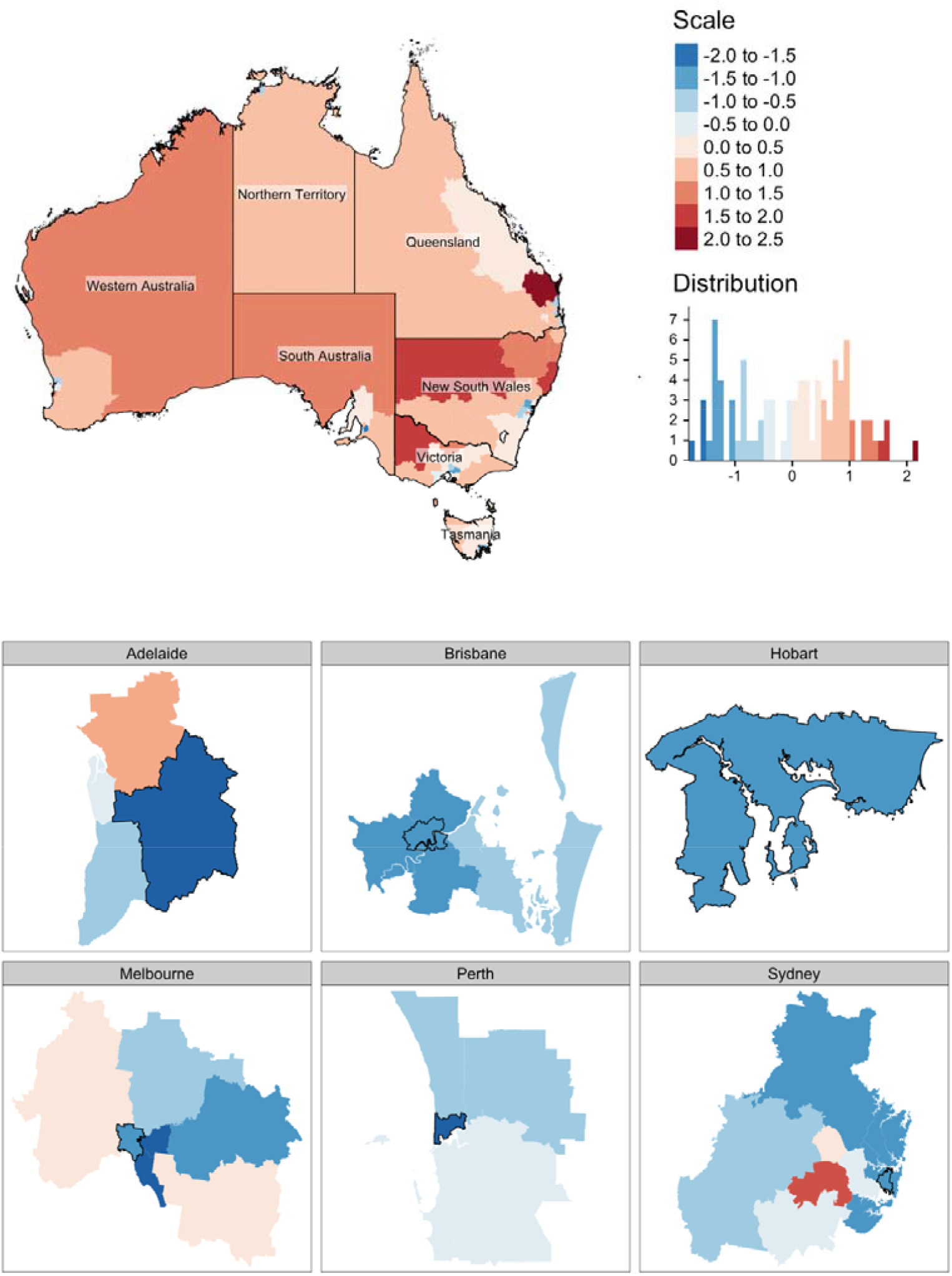
Mapping of scaled proportion of 2011 disadvantaged quintiles in a Statistical Area, where blue denotes below state average and red above state average, nationally and by state capitals; Statistical Areas with a black border in city maps are designed central/inner areas

**Supplementary figure 2.**
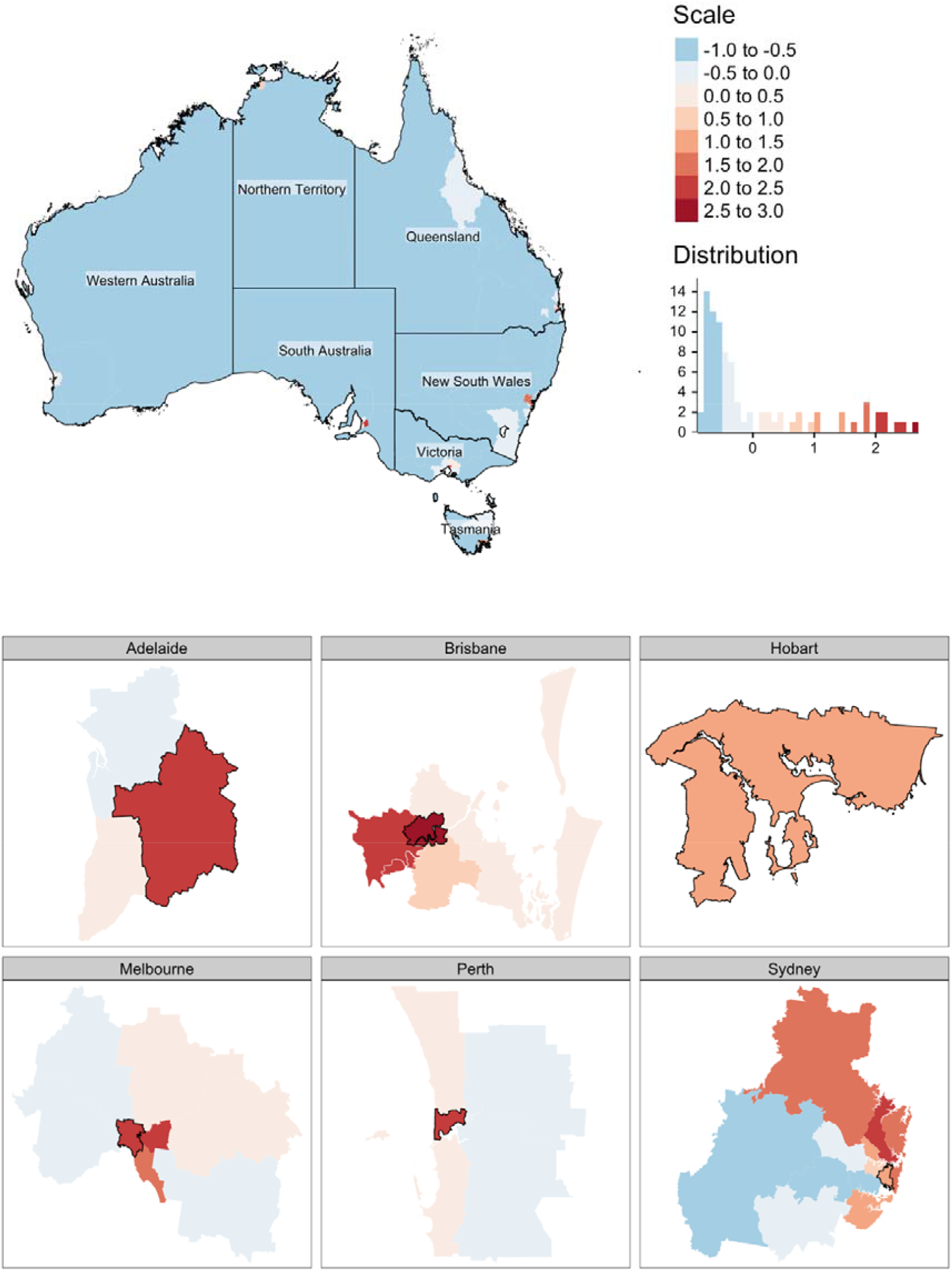
Mapping of scaled 2011 proportion of advantaged quintiles in a Statistical Area, where blue denotes below state average and red above state average, nationally and by state capitals; Statistical Areas with a black border in city maps are designed central/inner areas

**Supplementary figure 3.**
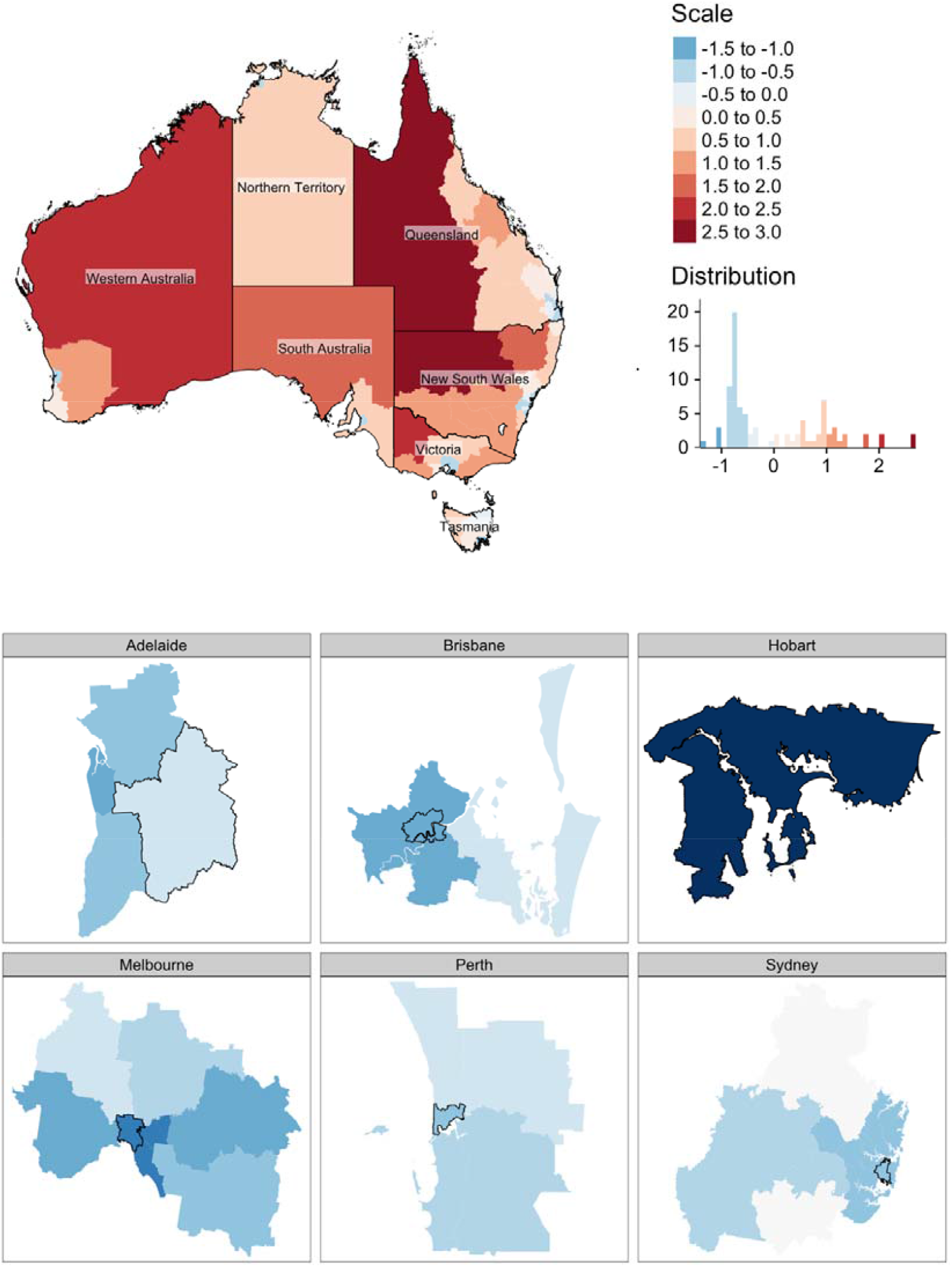
Mapping of scaled 2011 rurality score in a Statistical Area, where blue denotes below state average and red above state average, nationally and by state capitals; Statistical Areas with a black border in city maps are designed central/inner areas

**Supplementary figure 4.**
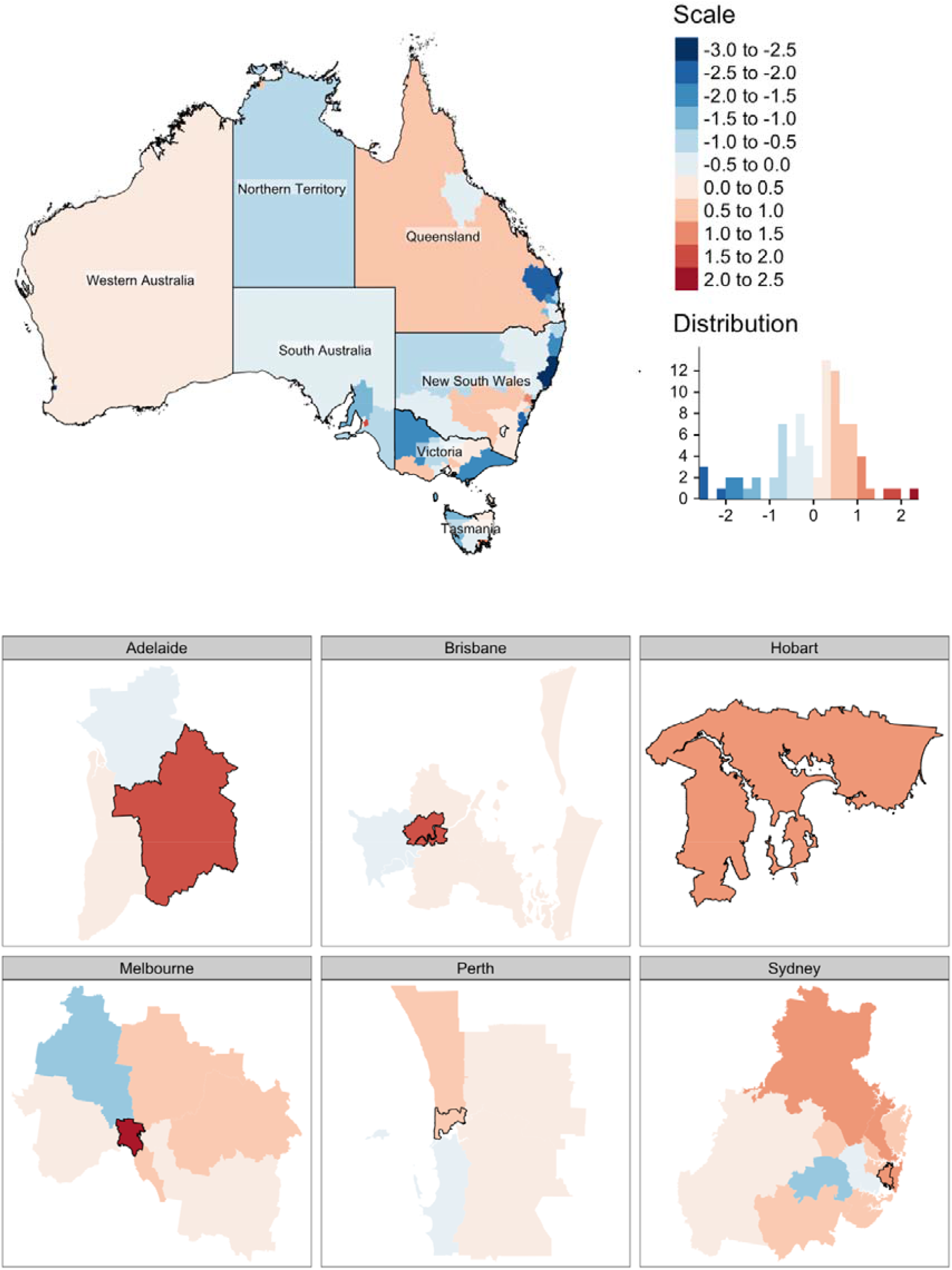
Mapping of scaled 2011 labour force participation rate in a Statistical Area, where blue denotes below state average and red above state average, nationally and by state capitals; Statistical Areas with a black border in city maps are designed central/inner areas

**Supplementary figure 5.**
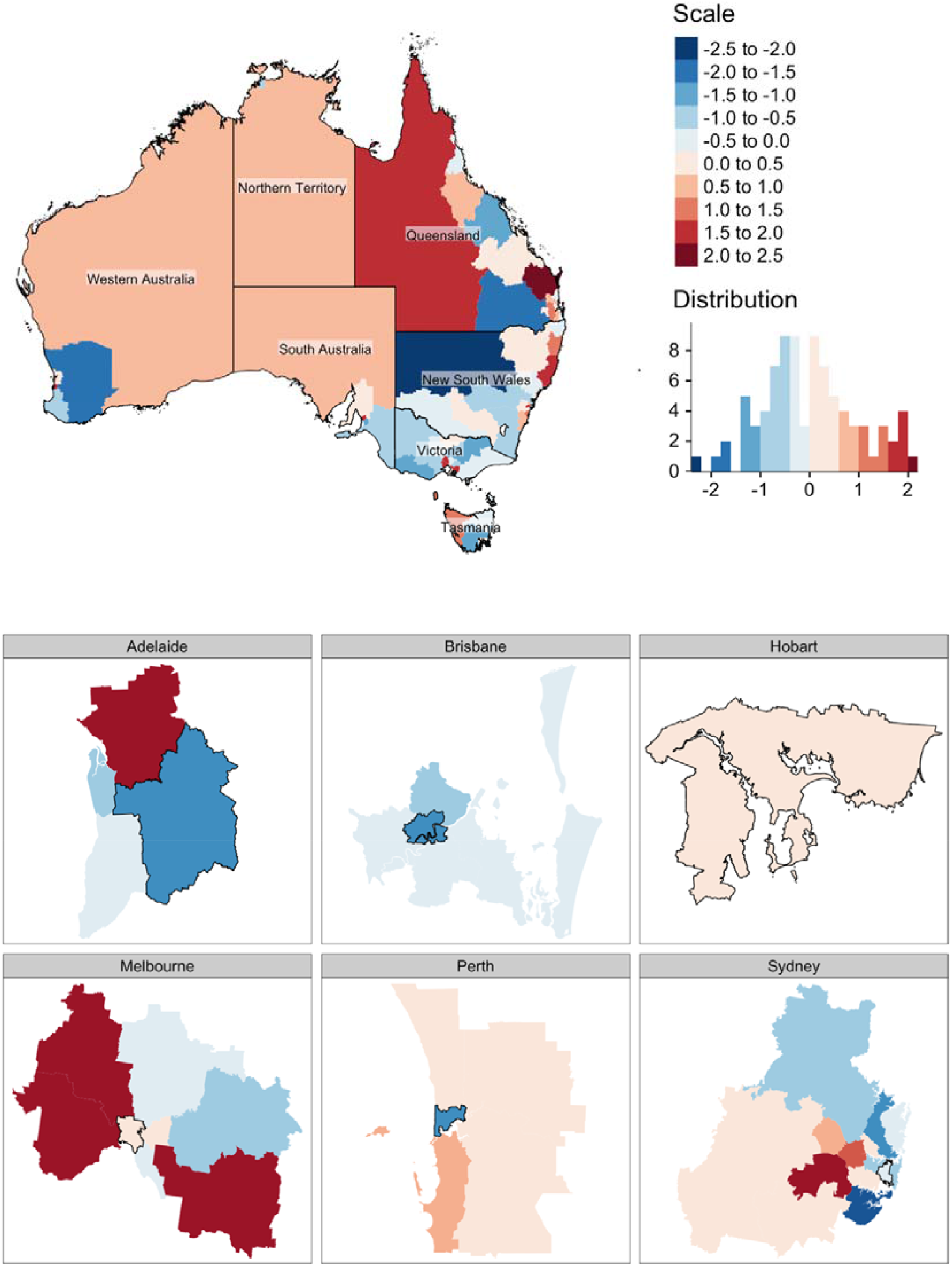
Mapping of scaled 2011 unemployment rate in a Statistical Area, where blue denotes below state average and red above state average, nationally and by state capitals; Statistical Areas with a black border in city maps are designed central/inner areas

**Supplementary figure 6.**
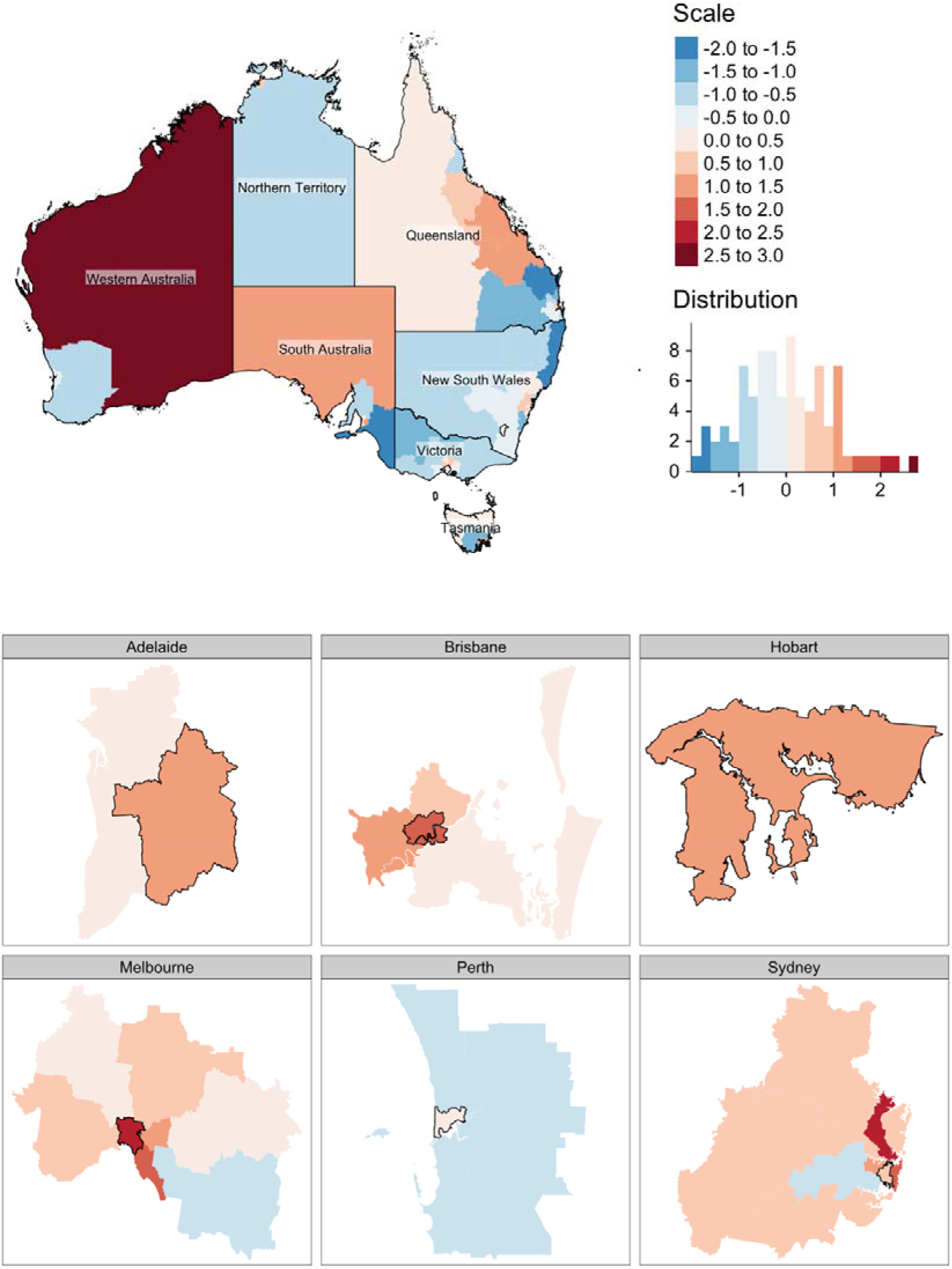
Mapping of scaled 2011 unemployment rate in a Statistical Area, where blue denotes below state average and red above state average, nationally and by state capitals; Statistical Areas with a black border in city maps are designed central/inner areas

**Supplementary figure 7.**
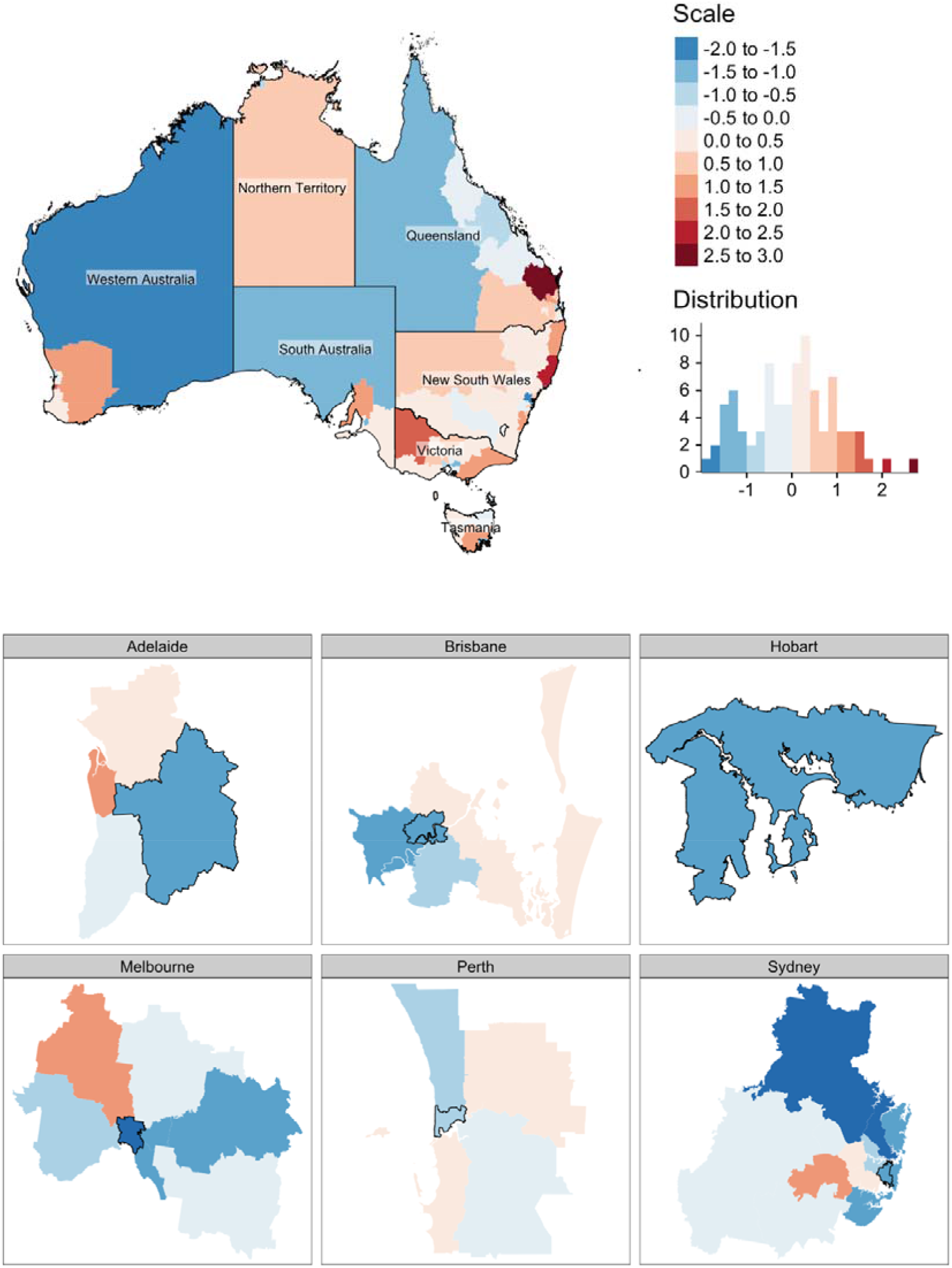
Mapping of scaled 2011 proportion of people needing assistance with core activities in a Statistical Area, where blue denotes below state average and red above state average, nationally and by state capitals; Statistical Areas with a black border in city maps are designed central/inner areas

**Supplementary figure 8.**
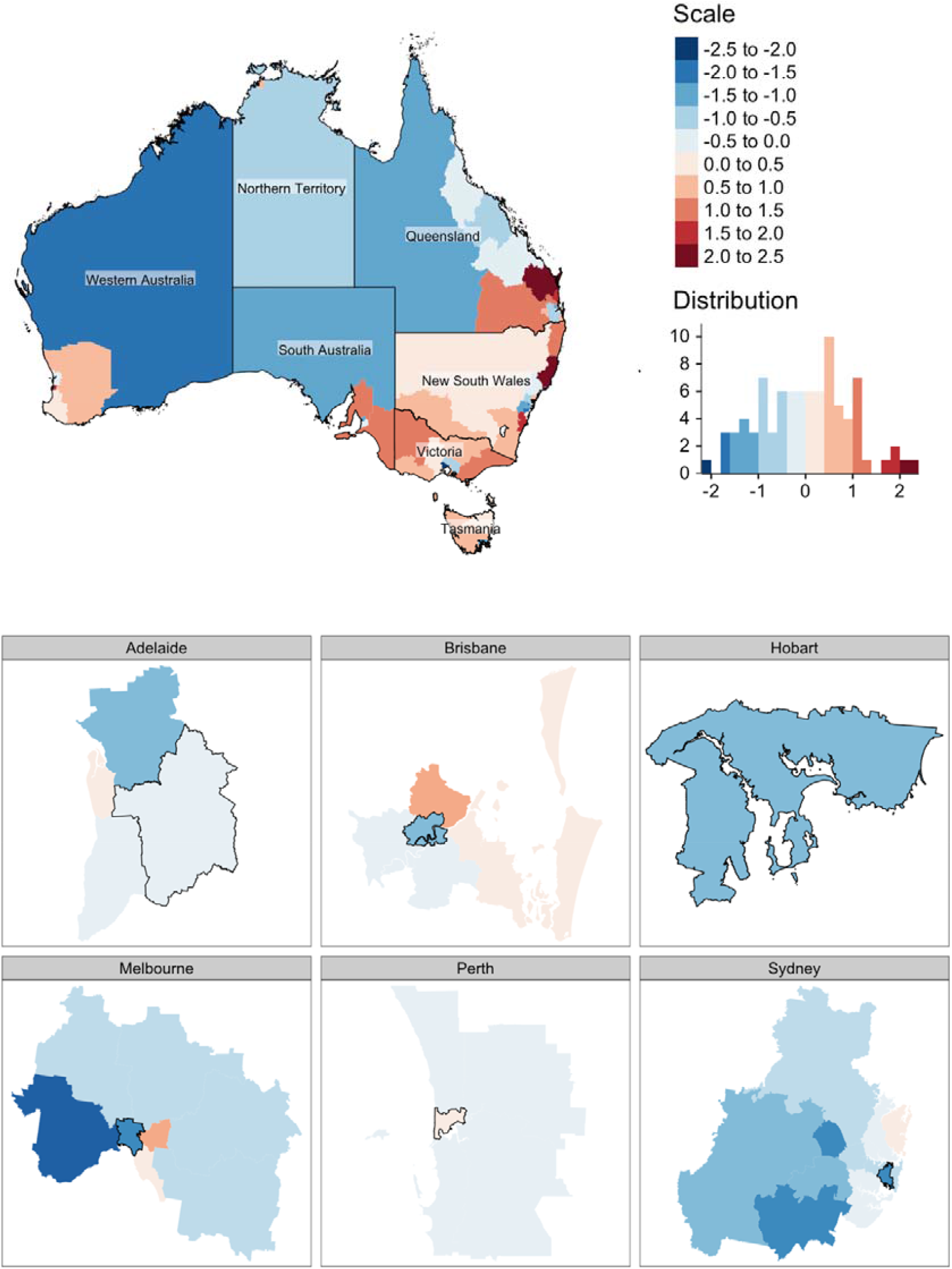
Mapping of scaled 2011 population aged 65+ years in a Statistical Area, where blue denotes below state average and red above state average, nationally and by state capitals; Statistical Areas with a black border in city maps are designed central/inner areas

**Supplementary figure 9.**
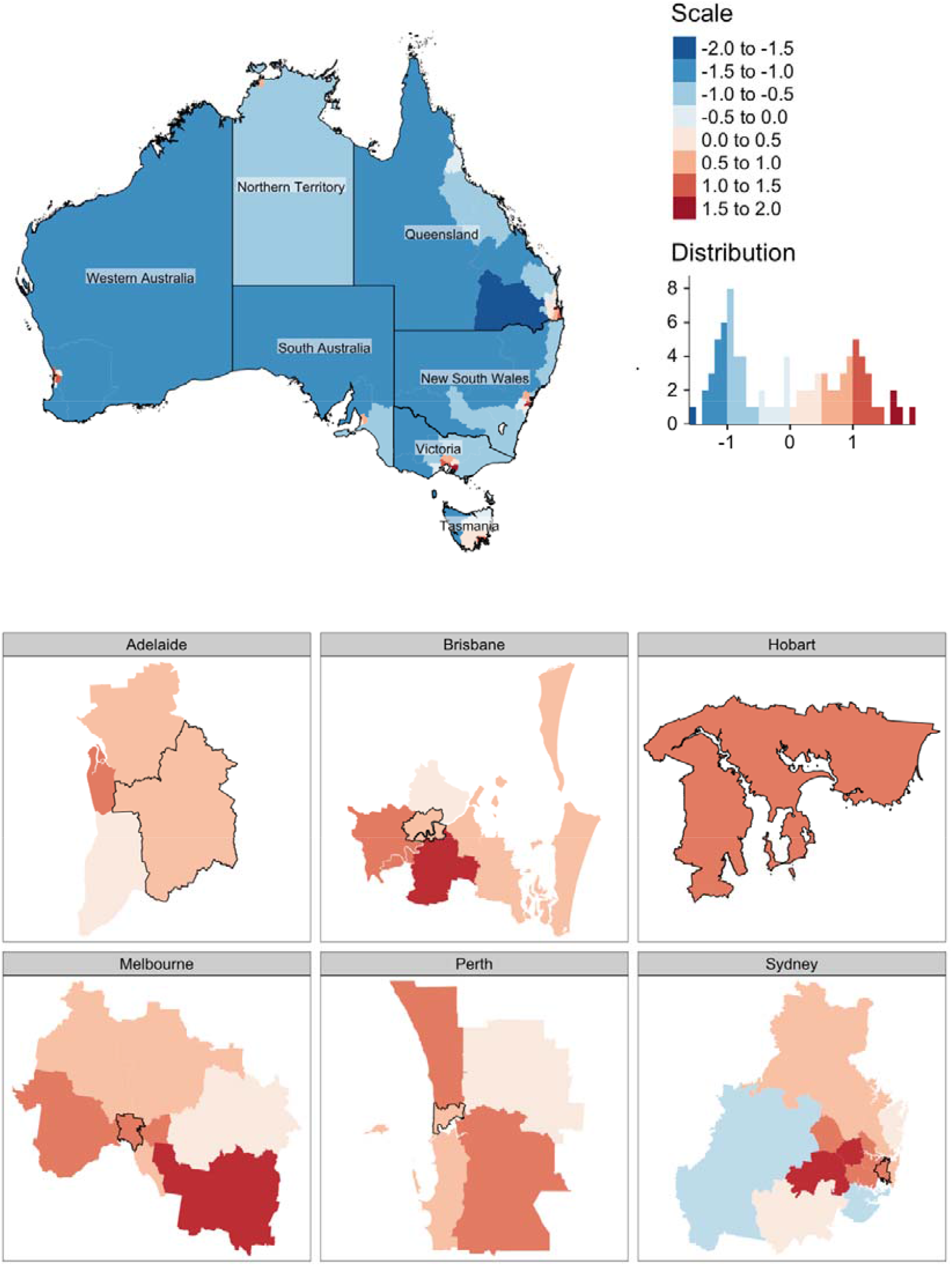
Mapping of scaled 2011 proportion of foreign-born in a Statistical Area, where blue denotes below state average and red above state average, nationally and by state capitals; Statistical Areas with a black border in city maps are designed central/inner areas

**Supplementary figure 10.**
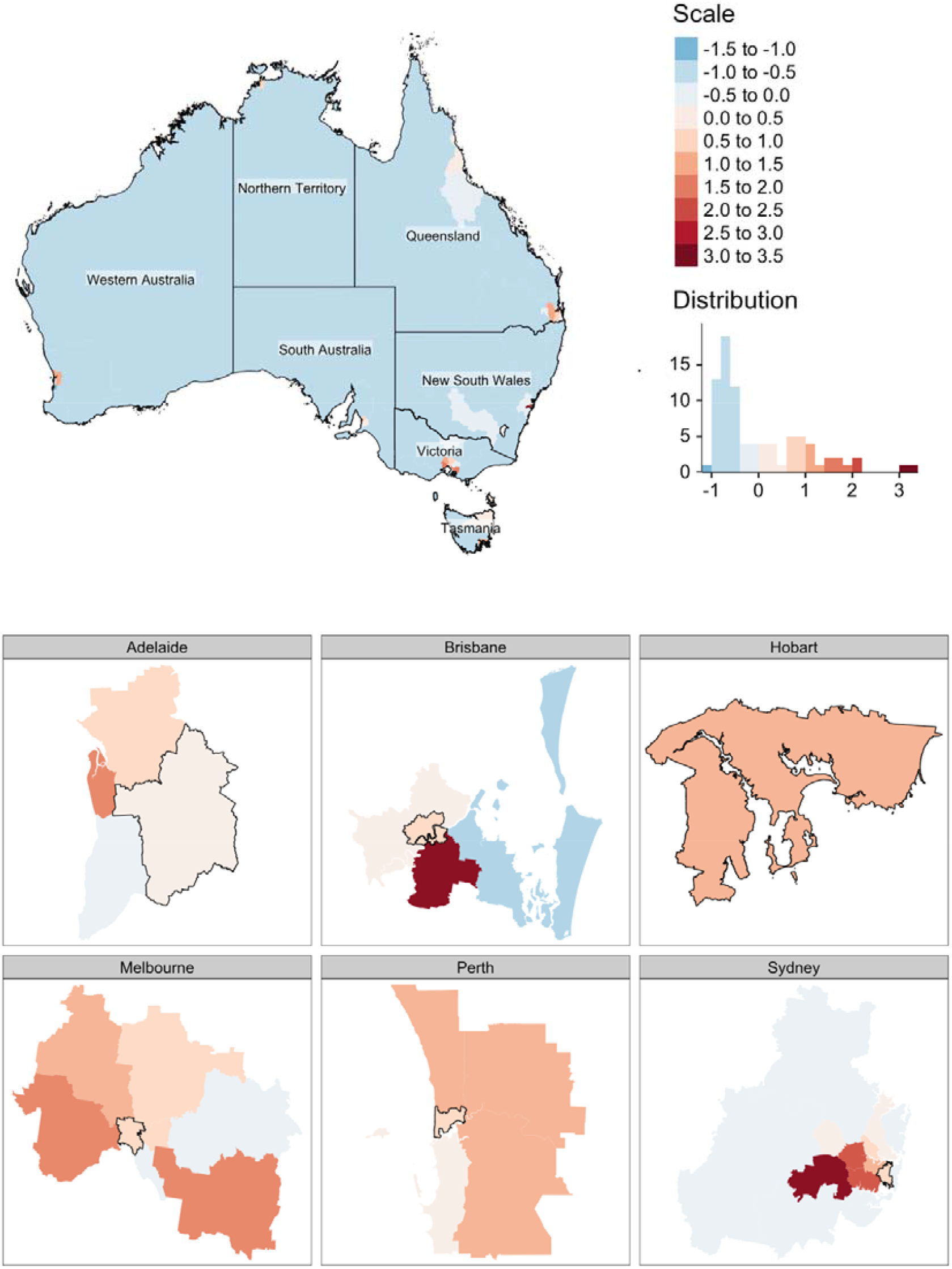
Mapping of scaled 2011 population with non-proficient English in a Statistical Area, where blue denotes below state average and red above state average, nationally and by state capitals; Statistical Areas with a black border in city maps are designed central/inner areas

**Supplementary figure 11.**
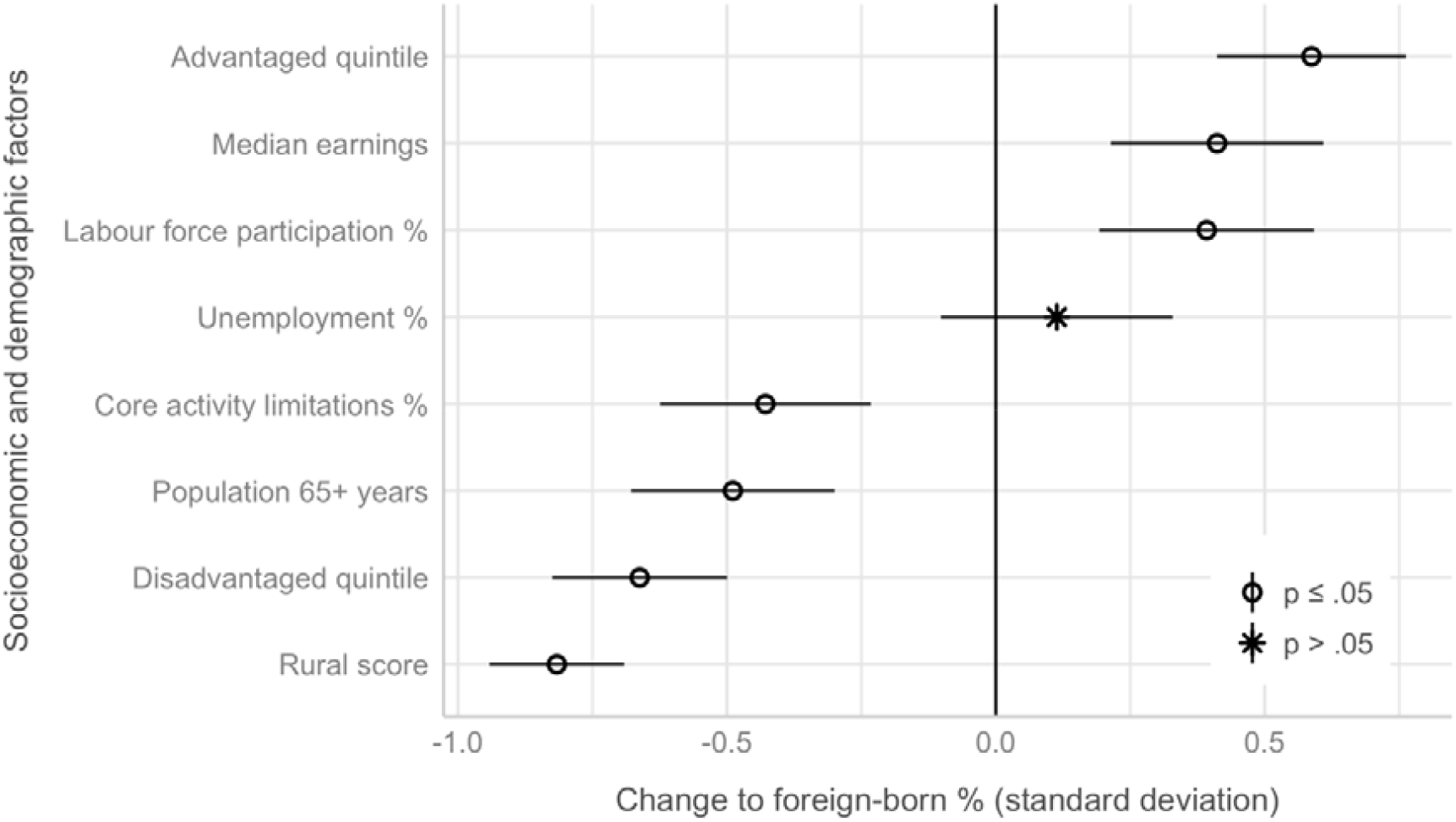
Association between proportion foreign-born and other sociodemographic factors (scores standardised)

**Supplementary figure 12.**
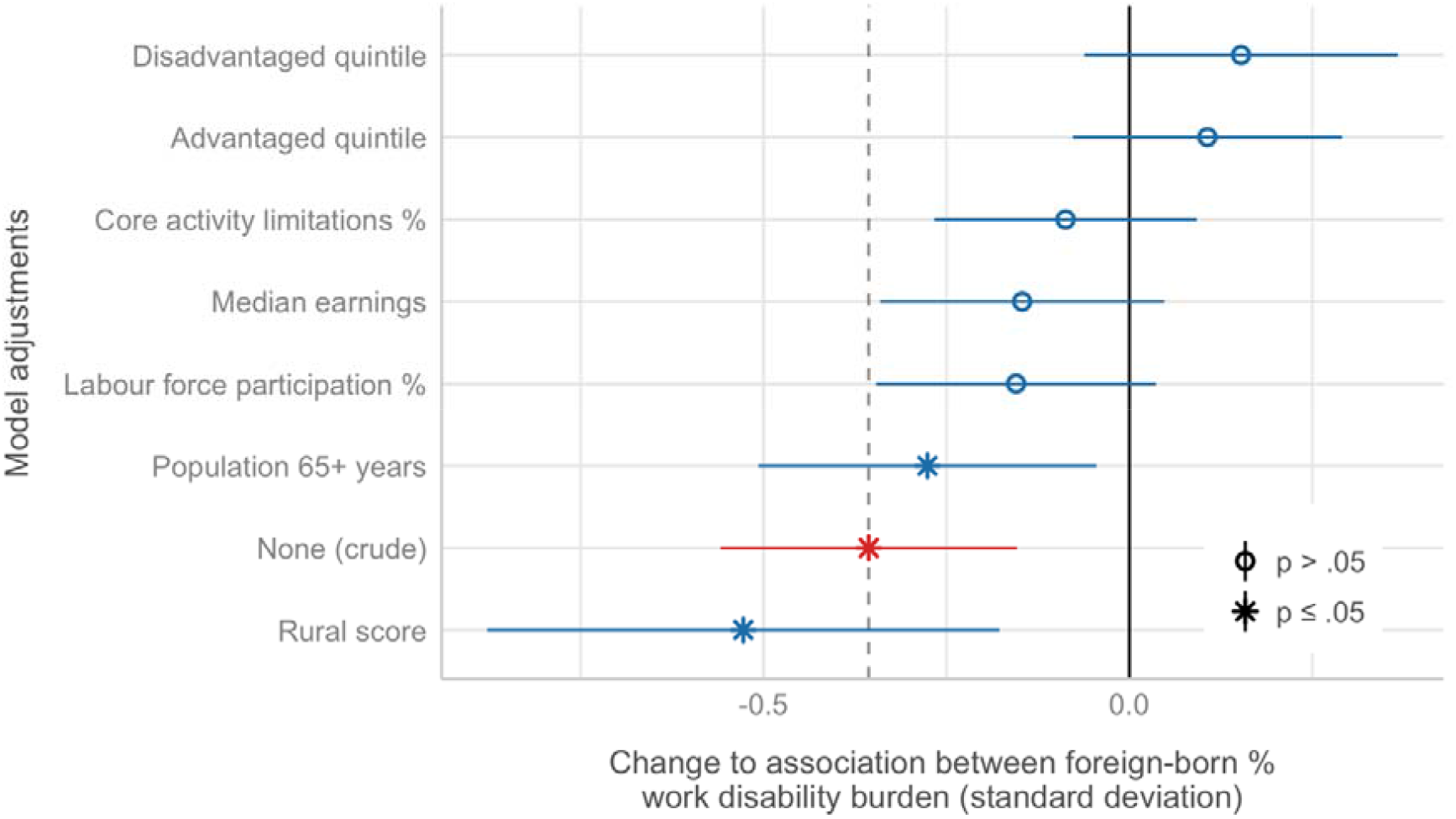
Change in association between proportion foreign-born and work disability burden when individually adjusting for sociodemographic factors that are significantly associated with both proportion of foreign-born and work disability burden (scores standardised)

## Notes

### Competing Interest Statement

The authors previously received salary support from funding provided by the workers compensation systems investigated in this study.

### Funding Statement

This study was funded by an Australian Research Council Discovery
Project Grant (DP190102473), as part of the Compensation and Return to Work Effectiveness (COMPARE) Project, and by Safe Work Australia, a government statutory agency that develops national work health and safety and workers compensation policy.

### Author Declarations

Monash Human Research Ethics Committee (CF14/2995 - 2014001663)

## REFERENCES

1. Collie A, Di Donato M, Iles RA. Work disability in Australia: an overview of prevalence, expenditure, support systems and services. J Occup Rehabil. 2019;29(3):526–539. doi:10.1007/s10926-018-9816-4

2. Costa-Black KM, Feuerstein M, Loisel P. Work Disability Models: Past and Present. In: Loisel P, Anema JR, eds. Handbook of Work Disability: Prevention and Management. Springer+Business Media; 2013:71–93. doi:10.1007/978-1-4614-6214-9

3. van Vreden C, Lane TJ, Iles RA, et al. Understanding the Impact of Place on Return to Work Following Work-Related Injury and Illness - a Scoping Review.

4. Gaines B, Besen E, Pransky GS. The influence of geographic variation in socio-cultural factors on length of work disability. Disabil Health J. 2017;10(2):308–319. doi:10.1016/j.dhjo.2016.12.009

5. Shraim M, Cifuentes M, Willetts JL, Marucci-Wellman HR, Pransky GS. Regional socioeconomic disparities in outcomes for workers with low back pain in the United States. Am J Ind Med. 2017;60(5):472–483. doi:10.1002/ajim.22712

6. Steenland K, Burnett C, Lalich N, Ward E, Hurrell J. Dying for work: The magnitude of us mortality from selected causes of death associated with occupation. Am J Ind Med. 2003;43(5):461–482. doi:10.1002/ajim.10216

7. Dembe AE, Delbos R, Erickson JB. The effect of occupation and industry on the injury risks from demanding work schedules. J Occup Environ Med. 2008;50(10):1185–1194. doi:10.1097/JOM.0b013e31817e7bf2

8. Young AE, Wasiak R, Webster BS, Shayne RGF. Urban-rural differences in work disability after an occupational injury. Scand J Work Environ Health. 2008;34(2):158–164.

9. Duckett S, Griffiths K. Perils of Place Identifying Hotspots of Health Inequality.; 2016. http://apo.org.au/files/Resource/874-perils-of-place.pdf

10. Wickizer TM, Franklin GM, Fulton-Kehoe D. Innovations in occupational health care delivery can prevent entry into permanent disability. Med Care. 2018;56(12):1018–1023. doi:10.1097/MLR.0000000000000991

11. Safe Work Australia. The Cost of Work-Related Injury and Illness for Australian Employers, Workers and the Community: 2012–13.; 2015.

12. Dembe AE. The social consequences of occupational injuries and illnesses. Am J Ind Med. 2001;40(4):403–417. doi:10.1002/ajim.1113

13. Collie A, Lane TJ. Australian workers’ compensation systems. In: Willis E, Reynolds L, Judge T, eds. Understanding the Australian Health Care System. 4th ed. Elsevier Australia; 2019:208–222.

14. Lippel K, Lötters F. Public Insurance Systems: A Comparison of Cause-Based and Disability-Based Income Support Systems. In: Loisel P, Anema JR, eds. Handbook of Work Disability. Springer; 2013:183–202. doi:10.1007/978-1-4614-6214-9

15. Lane TJ, Collie A, Hassani-Mahmooei B. Work-Related Injury and Illness in Australia, 2004 to 2014.; 2016.

16. Safe Work Australia. National Data Set for Compensation-Based Statistics Third Edition.; 2004.

17. Gray SE, Collie A. The nature and burden of occupational injury among first responder occupations: A retrospective cohort study in Australian workers. Injury. 2017;48:2470–2477. doi:10.1016/j.injury.2017.09.019

18. Krause N, Dasinger LK, Deegan LJ, Brand RJ, Rudolph L. Alternative approaches for measuring duration of work disability after low back injury based on administrative workers’ compensation data. Am J Ind Med. 1999;35(6):604–618. doi:10.1002/(SICI)1097-0274(199906)35:6<604::AID-AJIM8>3.0.CO;2-T

19. Australian Government. Employment Region Data. Labour Market Information Portal. Published 2020. Accessed July 29, 2020. https://lmip.gov.au/default.aspx?LMIP/Downloads/EmploymentRegion

20. Australian Bureau of Statistics. 1270.0.55.001 - Australian Statistical Geography Standard (ASGS): Volume 1 - Main Structure and Greater Capital City Statistical Areas, July 2016. Published 2016. Accessed July 30, 2020. https://www.abs.gov.au/ausstats/abs@.nsf/Lookup/bySubject/1270.0.55.001∼July2016∼MainFeatures∼Statistical Area Level 4 (SA4)∼10016

21. Australian Bureau of Statistics. 6524.0.55.002 - Personal Income in Australia, 2011-12 to 2016-17. Published 2019. Accessed July 30, 2020. https://www.abs.gov.au/AUSSTATS/abs@.nsf/Lookup/6524.0.55.002Main+Features12011-12to2016-17?OpenDocument

22. Australian Bureau of Statistics. 1410.0 - Data by Region, 2014-19. Published 2020. Accessed September 3, 2020. https://www.abs.gov.au/ausstats/abs@.nsf/mf/1410.0

23. ABS.Stat beta. ERP by SA2 and above (ASGS 2016), 2001 onwards. Published 2020. Accessed June 19, 2020. http://stat.data.abs.gov.au/Index.aspx?DataSetCode=ABS_ANNUAL_ERP_ASGS2016

24. Australian Bureau of Statistics. 1270.0.55.006 - Australian Statistical Geography Standard (ASGS): Correspondences, July 2011. Published 2012. https://www.abs.gov.au/AUSSTATS/abs@.nsf/DetailsPage/1270.0.55.006July2011?OpenDocument

25. Australian Bureau of Statistics. Research Paper: Socio-Economic Index for Areas - getting a handle on individual diversity within areas. Cat No 1351055036. Published online 2011.

26. Collie A, Lane TJ, Hassani-Mahmooei B, Thompson J, McLeod CB. Does time off work after injury vary by jurisdiction? A comparative study of eight Australian workers’ compensation systems. BMJ Open. 2016;6(5):e010910. doi:10.1136/bmjopen-2015-010910

27. Lane TJ, Sheehan LR, Gray SE, Collie A. Regional Differences in Time off Work after Injury: A Comparison of Australian States and Territories within a Single Workers’ Compensation System.; 2020. doi:10.1101/2020.07.23.20160416

28. Australian Bureau of Statistics. 2071.0 - Census of Population and Housing: Reflecting Australia - Stories from the Census, 2016. Published 2018. Accessed June 17, 2020. https://www.abs.gov.au/ausstats/abs@.nsf/Lookup/bySubject/2071.0∼2016∼MainFeatures∼Socio-EconomicAdvantageandDisadvantage∼123

29. Australian Bureau of Statistics. 3101.0 - Australian Demographic Statistics, Dec 2016.

30. R Core Team. R: A language and environment for statistical computing. Published online 2020. https://www.r-project.org/

31. RStudio Team. RStudio: Integrated Development for R. Published online 2020. https://www.rstudio.com

32. Bolker B, Robinson D. broom.mixed: Tidying Methods for Mixed Models. Published online 2020.

33. Firke S. janitor: Simple Tools for Examining and Cleaning Dirty Data. Published online 2020.

34. Grolemund G, Wickham H. Dates and Times Made Easy with lubridate. J Stat Softw. 2011;40(3):1–25. doi:10.18637/jss.v040.i03

35. Bache SM, Wickham H. magrittr: A Forward-Pipe Operator for R. Published online 2014.

36. Wickham H, Bryan J. readxl: Read Excel Files. Published online 2019. https://cran.r-project.org/package=readxl

37. Lüdecke D, Makowski D, Waggoner P, Ben-Shachar MS. see: Visualisation Toolbox for “easystats” and Extra Geoms, Themes, and Color Palettes for “ggplot2”. Published online 2019. https://cran.r-project.org/package=see

38. Pebesma E. Simple Features for R: Standardized Support for Spatial Vector Data. R J. 2018;10(1):439–446. doi:10.32614/RJ-2018-009

39. Wickham H, Averick M, Bryan J, et al. Welcome to the Tidyverse Tidyverse package. J Open Source Softw.2019;4(43):1686. doi:10.21105/joss.01686

40. Tennekes M. tmap: Thematic Maps in R. J Stat Software2. 2018;84(6):1–39. doi:10.18637/jss.v084.i06

41. Zeileis A, Grothendieck G. zoo: S3 Infrastructure for Regular and Irregular Time Series. J Stat Softw. 2005;14(6):1–27. doi:10.1017/CBO9781107415324.004

42. Lane TJ, Collie A. Hotspots of work disability study: aggregated data and code. Bridges. doi:10.26180/5f7f8f8f5df60

43. Piha K, Laaksonen M, Martikainen P, Rahkonen O, Lahelma E. Socio-economic and occupational determinants of work injury absence. Eur J Public Health. 2013;23(4):693–698. doi:10.1093/eurpub/cks162

44. Fujishiro K, Xu J, Gong F. What does “occupation” represent as an indicator of socioeconomic status?: Exploring occupational prestige and health. Soc Sci Med. 2010;71(12):2100–2107. doi:10.1016/j.socscimed.2010.09.026

45. Oh JH, Shin EH. Inequalities in nonfatal work injury: The significance of race, human capital, and occupations. Soc Sci Med. 2003;57(11):2173–2182. doi:10.1016/S0277-9536(03)00073-X

46. Kennedy BP, Kawachi I, Glass R, Prothrow-Stith D. Income distribution, socioeconomic status, and self rated health in US (multiple letters) [6]. Br Med J. 1999;318(7195):1417–1418. doi:10.1136/bmj.318.7195.1417a

47. Buist-Bouwman MA, De Graaf R, Vollebergh WAM, Ormel J. Comorbidity of physical and mental disorders and the effect on work-loss days. Acta Psychiatr Scand. 2005;111(6):436–443. doi:10.1111/j.1600-0447.2005.00513.x

48. Australian Institute of Health and Welfare. Rural & Remote Health.; 2019. https://www.aihw.gov.au/reports/rural-remote-australians/rural-remote-health

49. Matz CJ, Stieb DM, Brion O. Urban-rural differences in daily time-activity patterns, occupational activity and housing characteristics. Environ Heal A Glob Access Sci Source. 2015;14(1):1–11. doi:10.1186/s12940-015-0075-y

50. Lane TJ, Gray SE, Sheehan LR, Collie A. Increased benefit generosity and the impact on workers’ compensation claiming behavior: an interrupted time series study in Victoria, Australia. J Occup Environ Med. 2019;61(3):e82–e90. doi:10.1097/jom.0000000000001531

51. Hansen B, Nguyen T, Waddell GR. Benefit Generosity and Injury Duration: Quasi-Experimental Evidence from Regression Kinks.; 2017. http://ftp.iza.org/dp10621.pdf

52. Safe Work Australia. Comparison of Workers’ Compensation Arrangements in Australia and New Zealand.; 2020. https://www.safeworkaustralia.gov.au/doc/comparison-workers-compensation-arrangements-australia-and-new-zealand-2019

53. Reid A, Lenguerrand E, Santos I, et al. Taking risks and survival jobs: Foreign-born workers and work-related injuries in Australia. Saf Sci. 2014;70(March 2013):378–386. doi:10.1016/j.ssci.2014.07.002

54. Australian Bureau of Statistics. 6324.0 Work-Related Injuries, Australia, Jul 2017 to Jun 2018. Published 2018. AccessedMay 7, 2020 https://www.abs.gov.au/ausstats/abs@.nsf/mf/6324.0.

